# CNN and LSTM Models for fMRI-based Schizophrenia Classification Using c-ICA of dFNC

**DOI:** 10.1101/2025.02.27.25322899

**Authors:** Moein Esfahani, Hemanth Venkateswara, Zening Fu, Ram Ballem, Vince Calhoun

**Affiliations:** Department of Computer Science, Georgia State University, Atlanta, GA

**Keywords:** rs-fMRI, ICA, Schizophrenia, CNN, LSTM, Classification

## Abstract

Resting-state fMRI (rs-fMRI) captures brain activity at rest, it demonstrates information on how different regions interact without explicity task-based influences. This provides insights into both healthy and disordered brain states. However, clinical application of rs-fMRI remains challenging due to the wide variability in functional connectivity across individuals. Traditional data-driven methods like independent component analysis (ICA) struggle to balance these individual differences with broader patterns. Constrained methods, such as constrained ICA (cICA), have been introduced to address this by integrating templates from multiple external datasets to enhance accuracy and consistency. In our study, we analyzed rs-fMRI data from 100,517 individuals from diverse datasets, processed through a robust quality-control dynamic connectivity pipeline established in previous work. Using the resulting brain state templates as cICA priors, we examined the effectiveness of cICA for schizophrenia classification using a combined CNN and LSTM architecture. Results showed stable classification accuracy (87.6% to 86.43%) for the CNN model, while the LSTM model performed less optimally, likely due to sequence processing, yet still yielded comparable results. These findings underscore the potential of group-informed methods and prior data templates in constrained dynamic ICA, offering improved reliability and clinical relevance in rs-fMRI analysis and advancing our understanding of brain function.

## 1. INTRODUCTION

Resting-state functional MRI (rs-fMRI) has significantly advanced our understanding of brain function in both healthy and disordered states. By assessing functional interactions across the brain via the blood-oxygenation-level-dependent (BOLD) signal, rs-fMRI provides valuable insights. Unlike task-based fMRI, which requires subjects to engage in specific tasks, rs-fMRI captures spontaneous neural activity during rest, highlighting patterns of intrinsic connectivity rather than task-related features [1, 2]. However, despite its strengths, rs-fMRI faces limitations in clinical applications and single-subject analysis [1, 3]. A primary goal in rs-fMRI research is to uncover consistent connectivity patterns to better understand brain organization. Nevertheless, determining the ground truth for these functional patterns remains challenging. Unlike structural MRI, which relies on clearly defined anatomical structures, rs-fMRI is focused on uncovering latent functional networks. Achieving comparable functional patterns across individuals has proven difficult due to significant variability in connectivity [2, 4, 5]. Iraji et al [2, 5] addressed this challenge by identifying replicable networks to construct intrinsic connectivity network (ICN) templates. Building on this, in our previous work, we extended their application to dynamic functional network connectivity (dFNC) patterns. Leveraging data from over 100,000 subjects, we derived replicable dynamic states and used them as prior knowledge to estimate individual subjects’ dFNC patterns for comparison[6, 7]. To assess the utility of our generalized approach, we employed convolutional neural network (CNN) and long short-term memory (LSTM) architectures to classify schizophrenia (SZ) and healthy control (HC) subjects based on the estimated dFNC states[8, 9]. Our pipeline, detailed in fig. 1, involves input data with ICA and subsequently constrained ICA (cICA) before being processed by the same architecture. While classification result gains were relatively small, the resulting constrained dFNC pipeline resulted in more reliable and generalizable outcomes in addition to providing additional insights into brain dynamics.

**Fig. 1.**
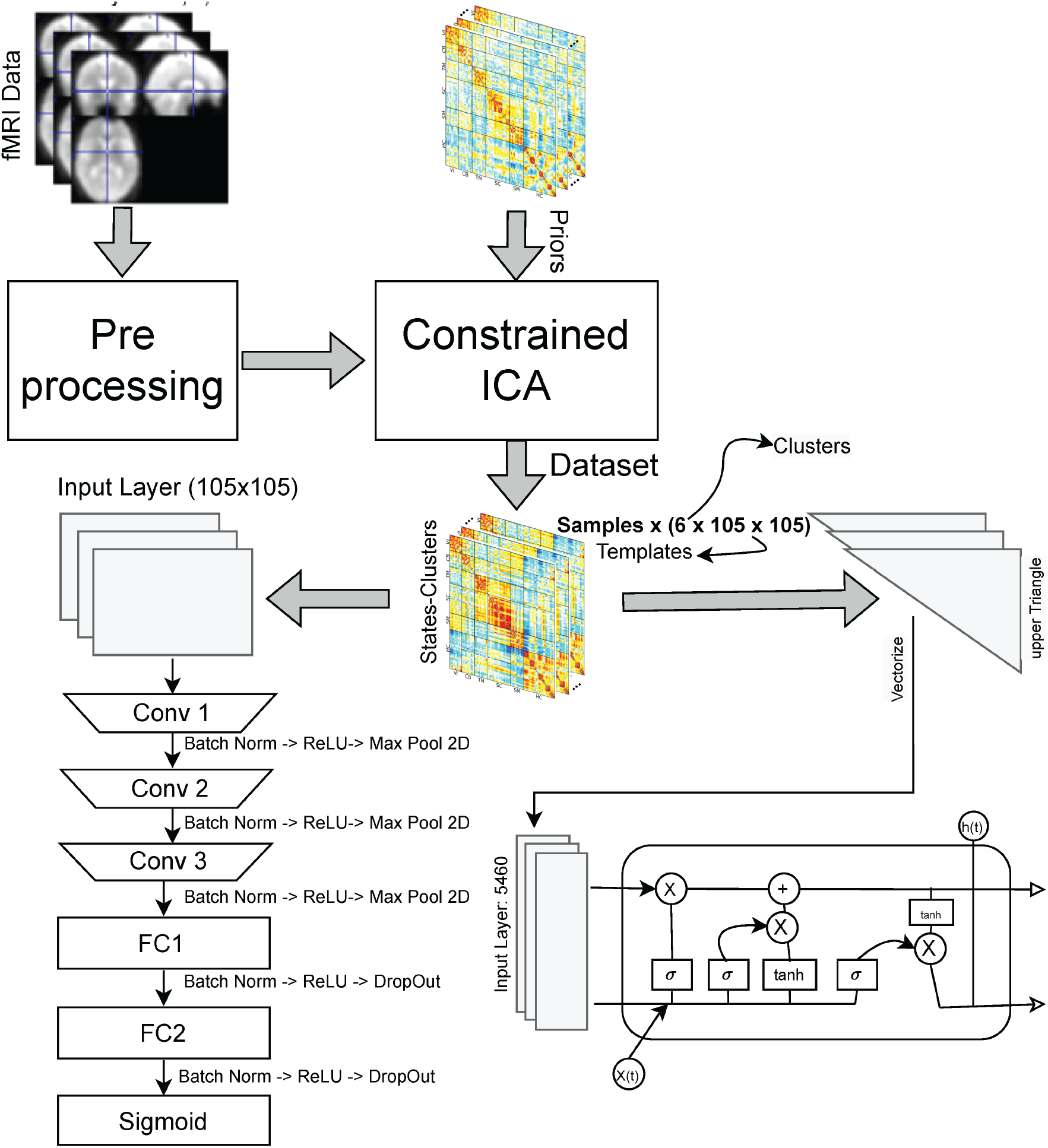
Schematic of the proposed pipeline, using templates as priors for constrained ICA. dataset were isolated and applied preprocessing, multiple deep learning models applied for classification, CNN behaved results as figures and LSTM behaves results as data sequences [for this figure I would highlight a couple states as well, the dynamics/ constrained dFNC is a major selling point for this work].

This paper is organized as follows: the introduction provides background on rs-fMRI and its challenges; materials and methods detail the datasets, preprocessing steps, and proposed deep learning architectures; the results and discussion examine classification outcomes; and the conclusion highlights future research directions for enhancing rs-fMRI clinical applications.

## 2. MATERIALS AND METHODS

### 2.1. Dataset

In this study, we analyzed resting-state fMRI data from release 2 of the multi-site Bipolar and Schizophrenia Network on Intermediate Phenotypes (BSNIP-2) study 1[10].also, for generalizability we aslso used some subjects of BSNIP1[11]. BSNIP-2 is the extension of BSNIP-1 data and was designed to repeat the same processes with enhanced proband number, biomarker panel, and sophistication of multivariate statistical approaches. BSNIP-2 includes six participated sites located in Athens, Boston, Chicago, Dallas, Hartford and Georgia, which recruited clinically stable, outpatient participants with diagnoses of schizophrenia (SZ), schizoaffective disorder (SAD), or bipolar disorder with psychosis (BDP) and healthy participants.for this study we have two main classes inclusing Schizophrenia (SZ) and healthy control (HC). Written consent was approved by each site’s respective Institutional Review Board. Participants completed the Structured Clinical Interview for the DSM-IV-TR. BSNIP-2 collected clinical, psychosocial, electrophysiological, imaging, and blood biomarkers for psychotic disorders. BSNIP-2 data included 1206 participants with both imaging sessions and clinical measurements available at the NIMH Data Archive (nda.nih.gov). All participants engaged in a focused 5-minute resting state fMRI scan and a detailed 3D T1-weighted structural scan on a 3T scanner at each site. Participants were carefully instructed to keep their eyes open, concentrate on a crosshair displayed on a monitor, and remain completely still throughout the process. Similar to BSNIP-1 study, BSNIP-2 confirmed alertness levels of each participant immediately following the scan, ensuring high-quality data collection[2, 12]. The fMRI data were preprocessed using statistical parametric mapping <u>(SPM12)</u> under MATLAB 2022b environment. We first discarded the initial 10 dummy volumes to allow the tissues reach steady state. A rigid body motion correction was performed using the toolbox in SPM to correct for subject head motion, followed by the slicetiming correction to account for timing difference in slice acquisition. The fMRI data were subsequently warped into the standard Montreal Neurological Institute (MNI) space using an echo planar imaging (EPI) template and were slightly resampled to 3 × 3 ×3 mm3 isotropic voxels. The resampled fMRI images were finally smoothed using a Gaussian kernel with a full width at half maximum (FWHM) = 6 <u>mm</u>[2, 12].

### 2.2. Convolutional Neural Network Model

In this study, we proposed and used two deep learning architectures, The convolutional neural network (CNN) model is designed to process multi layer input of size 6 × 105 × 105, this model designed specifically to capture the diffrences of this special data.note that we do not call these data as images and do not behave them as images, however we illustrated them before in other publications and in **fig**. This architecture employs three convolutional layers followed by two fully connected layers, with batch normalization, ReLU activations, max-pooling, and dropout to enhance performance and reduce overfitting. The final layer is a sigmoid activation function for binary *HC&SZ* classification.

- **Input Layer**: The input layer consists of data with dimensions 105 × 105 for each layer. This size was based on the data output of constraint ICA with prioirs for this case study.
- **Conv (Convolutional Layer)**:
  - Convolutional layers consists of *k*_1_ = 3 filters (kernels) of size 32, 64, 128 filters, which slide across the input image with a stride of *s* = 1 and padding of *p* = 1. This layer extracts basic features such as edges and simple textures, which form the foundational patterns for deeper layers.
  - **Batch Normalization and ReLU Activation**: Batch normalization stabilizes the learning process by normalizing the input to the activation function, while the ReLU activation function introduces non-linearity, allowing the network to learn complex patterns.
  - **Max Pooling 2D**: multiple max-pooling layers are also applied, reducing the spatial dimensions by half and helping in translation invariance, as well as reducing computation.

After the convolutional layers, the feature maps are flattened into a one-dimensional vector and passed to fully connected layers for classification.

- **FC (Fully Connected Layers)**:
  - these layers consists of *n*_1_ neurons in sequence of 128, 512, 128, 1 and is responsible for interpreting the high-level features extracted by the convolutional layers.
  - **Batch Normalization, ReLU Activation, and Dropout**: Batch normalization and ReLU activation are applied, followed by dropout *p* = 0.5) to avoid overfitting by randomly deactivating neurons during training. These FC layers learns non-linear combinations of the extracted features to make predictions.

**Output Layer**: The output layer uses a sigmoid activation function, which outputs a probability value between 0 and 1 for binary classification. This allows the model to classify the input image into one of two categories based on a threshold (e.g., 0.5).

### 2.3. Long Short-Term Memory (LSTM) Model

next step, we proposed model utilizes a long short-term memory (LSTM) architecture to find features in temporal dependencies in sequential data. LSTM networks are specifically designed to address the vanishing gradient problem. In this model, the LSTM cell processes an input sequence *x*(*t*) with a hidden state *h*(*t*), using a series of gating mechanisms to selectively retain relevant information.The LSTM cell uses three gating mechanisms including forget, input and output gates. to regulate the flow of information, allowing the network to retain or discard information selectively.By dynamically adjusting the memory cell state *c*_*t*_, the LSTM can learn which information to remember over long sequences.

## 3. RESULTS AND DISCUSSION

In this paper, based on other publications[2] we used the computed templates as priors for constrained ICA model.applying multiple ICA models to fMRI data doesnt necessarily change the results in accuracy model because the concept are not diffrent. Based on fig. 3, we have accuracy results for multiple training models. fig. 3 shows that LSTM model has poor performance in classification of these results in general. However, in fig. 2 the confusion matrices present the results of two deep learning models (CNN and LSTM), each tested with and without cICA. The matrices show true positives (TP), true negatives (TN), false positives (FP), and false negatives (FN) for two groups where each individual was either a schizophrenia (SZ) or a healthy control (HC). For the cICA with CNN Model, the configuration yields a high number of true positives (TP = 113) and true negatives (TN = 113), indicating strong performance in classifying both SZ and HC groups correctly. The false positives (FP = 13) and false negatives (FN = 19) are relatively low, suggesting a balanced accuracy across both classes. This model is effective at maintaining high precision and recall. for ICA with CNN Model,Without the constrained ICA, the CNN model has a slightly lower TP count (TP = 110) but retains the same TN value (TN = 113). As mentioned earlier, the performance is comparable but slightly reduced without the constrained ICA, showing that constrained ICA might contribute to a better balance between precision and recall. However, for the cICA with LSTM Model, the configuration achieves a TP of 82 and TN of 51, which are lower than those of the CNN model. Additionally, the FP and FN counts are higher, with FP = 75 and FN = 50. This suggests that the LSTM model struggles more with classification accuracy compared to the CNN model and has lower performance overall. For the ICA with LSTM Model With ICA but without priors, the LSTM model shows a slight improvement in TP (84) and TN (59) compared to the constrained ICA setting. However, the FP (67) and FN (48) rates are still high, though slightly reduced compared to the constrained ICA version. The results demonstrates several key insights into the effectiveness of constrained ICA and the impact of model choice (CNN vs. LSTM) on classification performance. Constrained ICA appears to enhance the CNN model’s performance by marginally increasing its TP and TN counts and lowering its FP and FN rates. This improvement suggests that constrained ICA may help reduce noise a bit better or magnifies key features that the CNN model uses effectively for classification. However, the impact of constrained ICA on the LSTM model is less favorable. The LSTM model shows lower accuracy with constrained ICA, possibly because it relies more on temporal dependencies. The CNN model consistently outperforms the LSTM model across all architectures. The CNN’s higher TP and TN values, coupled with lower FP and FN rates, demonstrate that it is better suited for this classification task, especially when constrained ICA is applied. The LSTM model’s performance, even with ICA, does not reach the same level as that of the CNN model. This may indicate that the CNN architecture is better suited for extracting spatial features, which are critical for distinguishing between SZ and HC groups in this dataset. In contrast, the LSTM model’s focus on sequential dependencies might be less relevant, leading to more classification

**Fig. 2.**
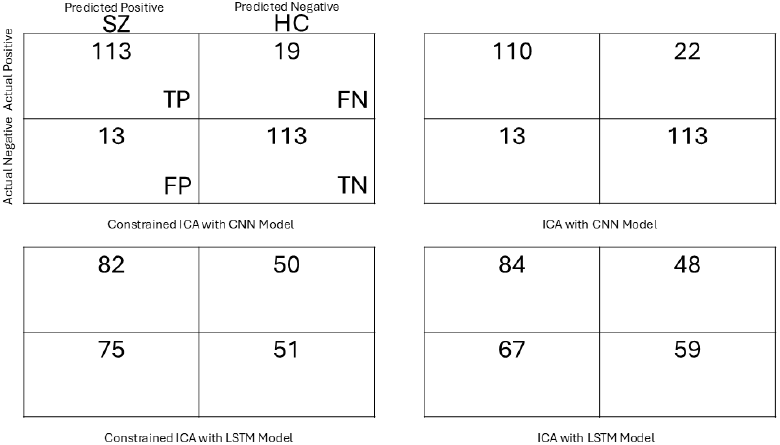
multiple case studies confusion matrix of the results for tests, including 258 subjects, 132 SZ and 126 HC classes.

**Fig. 3.**
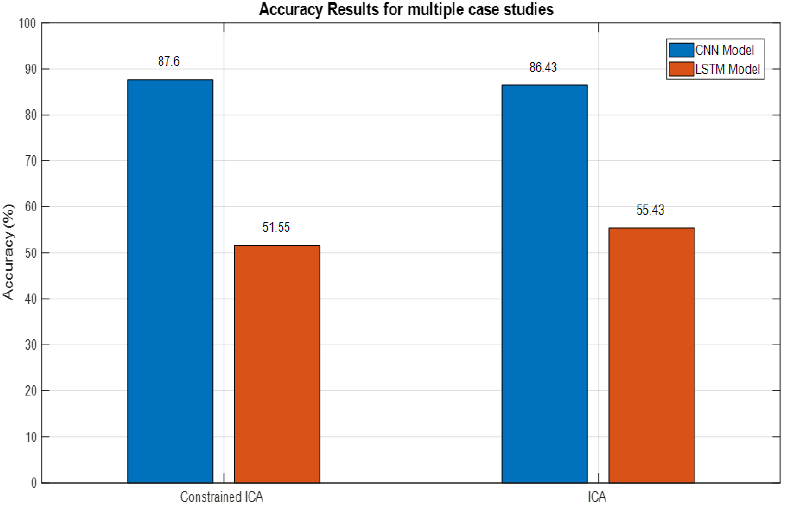
Accuracy compression of multiple models with Constrained ICA vs ICA *withoutpriors* errors.

## 4. CONCLUSION

In this study, we proposed a pipeline for classification of schizophrenia using CNN and LSTM models applied to rs-fMRI data, we then used constrained ICA and then constrained dFNCs for classification. By using data-driven templates from large external datasets, the priors and templates helped us resulting in more consistent and reliable patterns of dFNC across subjects. Our findings demonstrated that the CNN model maintained high classification accuracy when constrained ICA templates were applied, while the LSTM model, though challenged by sequence processing, produced comparable results. This suggests that cICA effectively stabilizes classification performance by using priors. Overall, our results highlight the potential of group-informed methods like cICA and cinstrained dFNC to improve generalizability and clinical relevance in neuroimaging applications, particularly in the context of mental health conditions such as schizophrenia. Future research could expand on this foundation by tuning these architectures, exploring additional clinical conditions and multiple datasets, and enhancing template-based approaches like time frequency moments to further improve accuracy and robustness in rs-fMRI classification tasks.

## Data Availability

dataset is only available by NIH archive upon request

https://www.nih.gov/

## 5. ACKNOWLEDGMENTS

This work was supported by grants from the NIH (R01MH123610) and NSF (2316421)

